# Safety and adherence to self-administered intravaginal 5-fluorouracil cream following cervical intraepithelial neoplasia (CIN) 2/3 treatment among HIV-positive women in Kenya: A phase 1 clinical trial

**DOI:** 10.1101/2024.06.06.24308548

**Authors:** Chemtai Mungo, Jackton Omoto, Cirilus Ogollah, Antony Owaya, Mercy Rop, Gershon Rota, Elizabeth A Bukusi, Jennifer Tang, Lisa Rahangdale

## Abstract

**Objective:** To determine the safety, tolerance, and adherence to self-administered intravaginal 5% fluorouracil (5FU) cream as adjuvant therapy following cervical intraepithelial neoplasia grade 2 or 3 (CIN2/3) treatment among women living with HIV (WLWH) in Kenya.

**Methods:** A Phase I Pilot trial was performed among 12 WLWH in Kenya, aged 18–49 years between March 2023-February 2024 (Clinicaltrials.gov NCT05362955). Participants self-administered 2g of 5FU intravaginally every other week for eight applications. Safety was assessed using a standardized grading scale, and adherence was evaluated using self-report, inspection of used applicators, and weighing of the study drug.

**Results:** The mean age and CD4 count were 43.9 years and 781 cells/mm^3^, respectively. Seven (58%) had an 8^th^-grade education or less. All 12 reported at least one grade I adverse event (AE), 1 (8%) reported a grade 2 AE, no grade 3 or 4 AEs were reported. Increased vaginal discharge (n=9, 75%) and irritation (n=5, 42%), with a mean duration of 3.2 and 2.8 days, respectively, were the most commonly reported AEs. Provider-observed AEs included grade 1 cervical erythema and superficial abrasions. All participants tolerated all eight 5FU doses, and 96% adherence was demonstrated.

**Conclusion:** Self-administered 5FU following CIN2/3 treatment among WLWH in Kisumu, Kenya, was safe, tolerable, and associated with high adherence. Randomized trials are needed to investigate whether adjuvant 5FU can improve treatment outcomes or serve as primary cervical precancer treatment in sub-Saharan Africa. A self-administered therapy may be transformative in increasing access to treatment and, hence, secondary prevention of cervical cancer.

## INTRODUCTION

Although cervical cancer is preventable, in 2020, an estimated 604,000 new cases occurred, with low- and middle-income countries (LMICs) accounting for 85% of incident cases and 90% of deaths.^1^ Cervical cancer is an AIDS-defining malignancy, and women living with HIV (WLWH), the majority of whom reside in LMICs, have a six-fold increased risk of developing invasive cancer,^2^ making prevention efforts among them a priority. The lack of widespread screening, coupled with accessible treatment of precancerous lesions in LMICs, accounts for a disproportionate cervical cancer burden. To achieve the WHO’s 2030 targets for cervical cancer elimination, including adequately treating 90% of those with precancer or cancer,^3^ it is crucial to improve treatment of precancerous lesions among WLWH.

Available precancer treatment methods in LMICs are ablation or excision. However, among WLWH, both treatments are limited by high rates of recurrence of cervical intraepithelial neoplasia grade 2 and 3 (CIN2/3) or cervical precancer. Randomized trials in South Africa and Kenya demonstrate 18-19% and 27-30% CIN2/3 recurrence following excision and cryotherapy among WLWH, respectively.^4,5^ Rates of CIN2/3 recurrence among WLWH following thermal ablation are 28%^6^ - 39.9%^6,7^ at 1-3 years after treatment. This calls for studies on innovative yet feasible and accessible strategies to improve outcomes in this population, including topical therapies, immunotherapy, or therapeutic vaccinations.

One such therapy is self-administered topical 5% fluorouracil (5FU), an antimetabolite and cytotoxic drug used for the treatment of a variety of precancerous and malignant skin conditions.^8^ In high-income settings (HICs), studies have demonstrated the tolerability and efficacy of topical 5FU for the treatment of genital warts,^9^ vulvar,^10^ and vaginal^11^ precancers. Additionally, randomized trials in HICs have demonstrated the safety and efficacy of self-administered, intravaginal 5FU cream for cervical precancer.^12,13^

No studies have evaluated the feasibility of repurposing topical 5FU - a generic therapy available in LMICs and on the WHO List of Essential Medications^14^ – for cervical precancer treatment in WLWH in LMICs. To address this gap and inform future efficacy trials, we carried out a Phase I pilot study to evaluate the safety, uptake, tolerability, and adherence to adjuvant, self-administered intravaginal 5FU among WLWH in Kenya after primary CIN2/3 treatment.

## MATERIALS AND METHODS

We conducted a prospective single-arm, phase 1 pilot study among 12 WLWH at Lumumba Sub-County Hospital in Kisumu, Kenya, between March 10, 2023, and February 15, 2024. The study was approved by the institutional review boards at the University of North Carolina at Chapel Hill (21-3265) and the Kenya Medical Research Institute (KEMRI/SERU/CMR/4555). All participants provided informed consent.

The study protocol is published^15^ and the study flow diagram is shown in Figure 1.^15^ Participants were WLWH aged 18–49 years, with biopsy-confirmed CIN2/3, within 4-12 weeks of primary treatment and agreed to use dual contraception if of childbearing age. Exclusion criteria included pregnancy or breastfeeding, use of high-dose steroids, or prior history of cancer. After informed consent, medical history and eligibility criteria, baseline cervical images, and specimens for sexually transmitted infections (STI) and CD4 count testing were obtained. Participants diagnosed with STIs were offered treatment before enrollment. Participants attended an enrollment visit within 28 days of the screening visit(s) and received detailed counseling in the participant’s preferred language, usually *Dholuo*, on how to self-administer 5FU as outlined in Box 1 using pictorial aids and a pelvic model. Study participants were instructed to self-administer 2 grams of 5% 5FU cream (Effudex, Bausch Health, USA) intravaginally using an applicator biweekly (every two weeks) at night for eight doses. Participants were given the option to self-administer the first dose of 5FU in the clinic and the rest at home. Literate participants recorded date of cream use and adverse events in a study diary, while a nurse called non-literate participants weekly for this information. 5FU use was postponed if it coincided with the participant’s menstrual cycle or in cases of vaginal and vulvar abrasions. Sexual abstinence was required for 48 hours after each 5FU application.

#### Box 1

**Instructions for self-administration of Intravaginal 5-Fluorouracil (5FU)^1^**

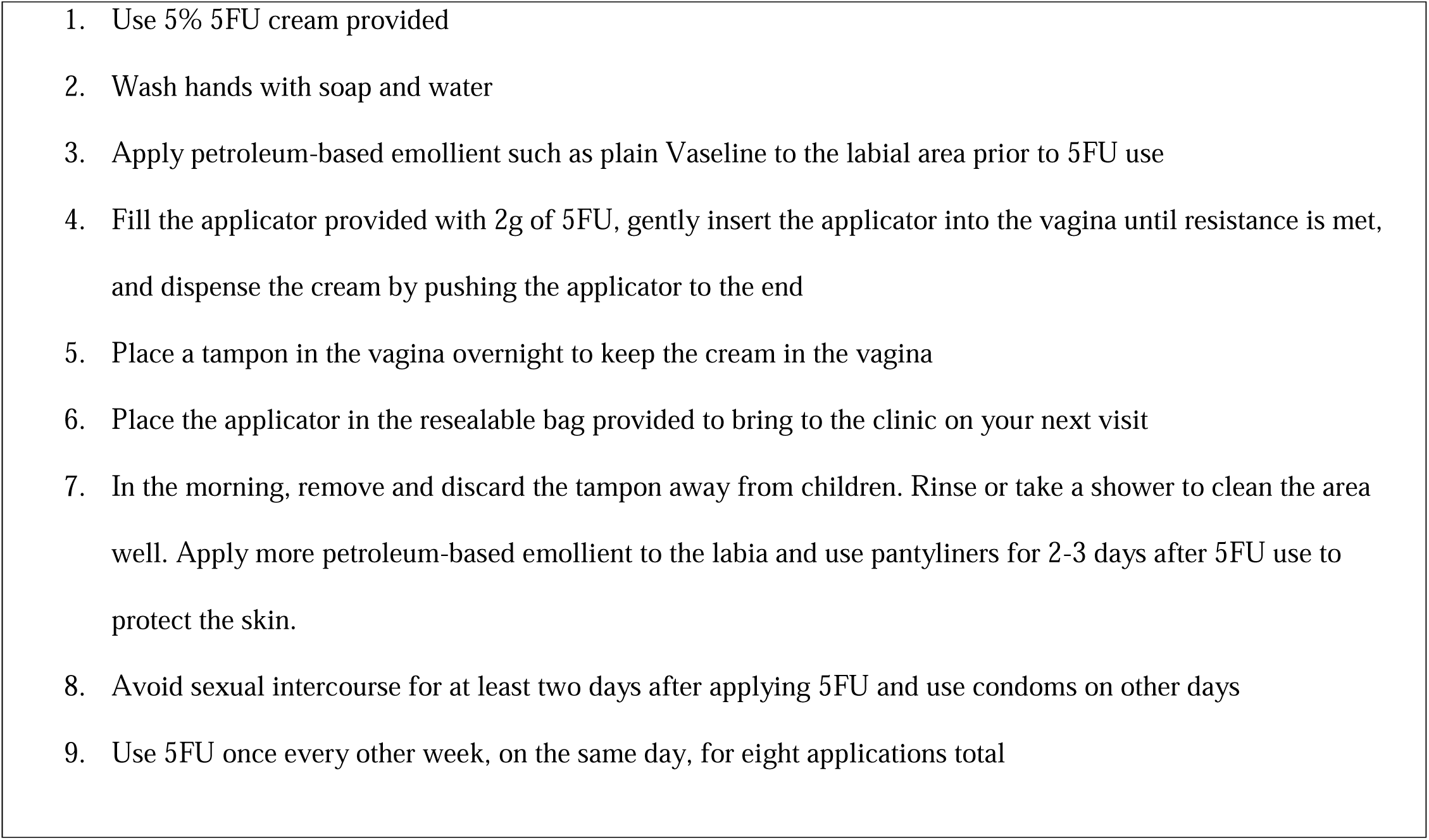

**Figure 1:**
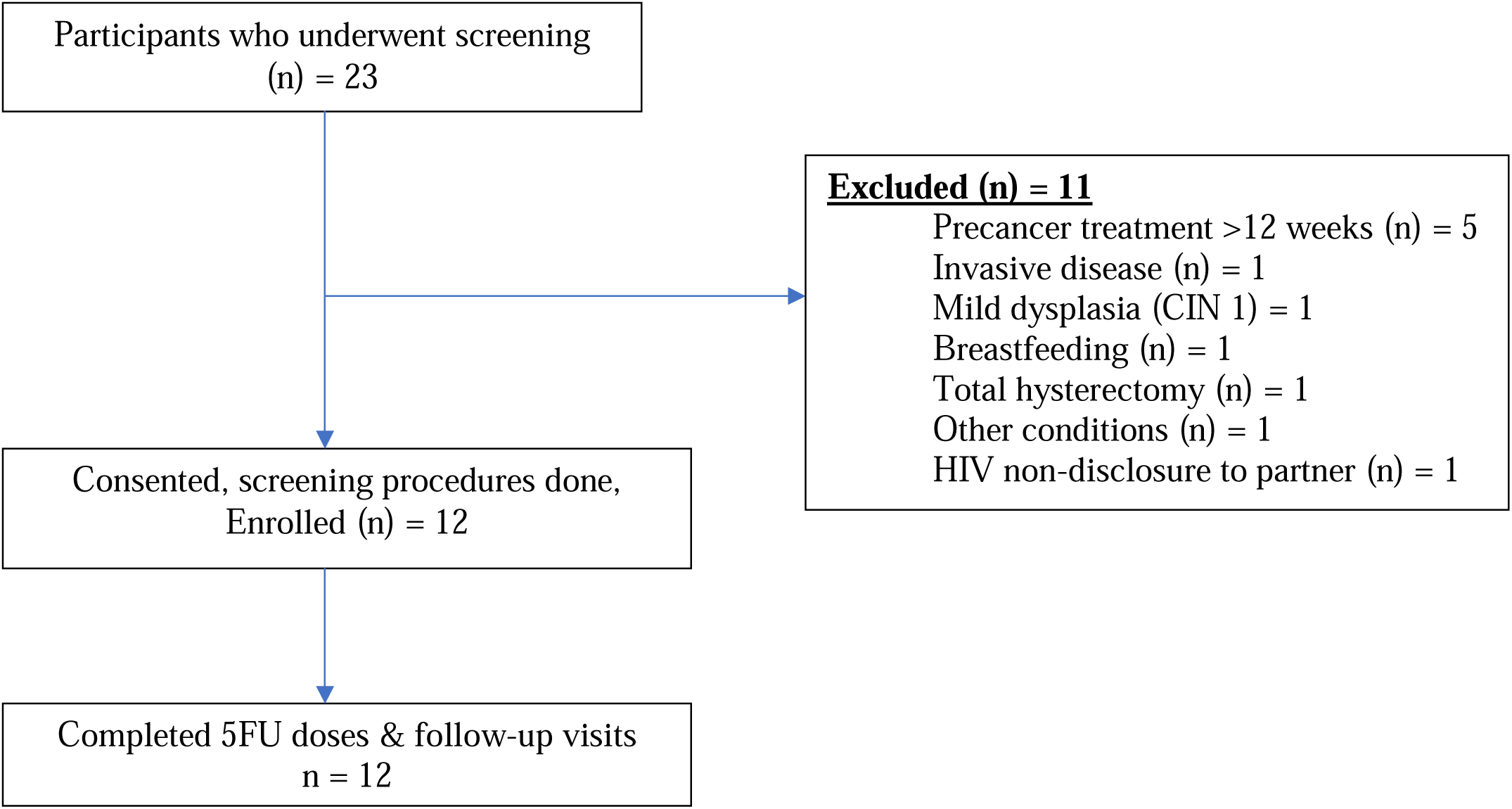
Flow diagram of participant recruitment into the 5FU Phase 1 trial in Kisumu, Kenya

Participants returned to the study clinic one week after each application for safety and adherence assessments and pregnancy testing. Adverse events (AEs) were reviewed from the study diaries, interviews, and pelvic examinations, and were graded using the standardized Division of AIDS Table for Grading the Adverse Events for Female Microbicide Trials.^16^ Three methods from previous microbicide trials ^17^ were used to evaluate adherence: 1) self-report (verbal or study diary), 2) weighing of study tubes at each visit to document appropriate decreasing weight, and 3) visual inspection of returned applicators for evidence of study drug (Supplemental Image 1). A participant was considered adherent to a specific 5FU dose if all three assessment methods confirmed its use. At week 16, acceptability was assessed through a questionnaire and an in-depth interview. Delayed adverse events were elicited at the final study visit on week 20. All participants were advised to follow up for repeat cervical cancer screening at 12 months after their primary treatment, as is the standard of care in Kenya.^18^

The primary endpoint of safety was evaluated by the type, frequency, severity, and duration of both reported and clinician-observed AEs during the trial, including 95% confidence intervals. The at-risk period for safety began at study Week 1 (after the first week of 5FU use) and continued through study Week 20 (4 weeks after the last 5FU use). Secondary endpoints were: 1) uptake, defined as the proportion of eligible screened participants who agreed to participate in the trial; 2) tolerability, defined as the proportion of participants who were unable or unwilling to apply at least 4 of 8 5FU doses due to adverse events or burden of self-application, 3) adherence. The study aimed to evaluate safety, specifically powered to exclude grade 3 or worse AEs, as is expected based on prior studies of intravaginal 5FU at this dose and frequency. With a sample size of 12 and assuming no grade 3 or worse AEs occur, the upper limit of the 95% confidence interval of the AE rate would be 26.5%.^19^ This means we can confidently rule out occurrence of grade 3 or worse AEs in 26.5% of the population.

All analyses were conducted using Stata version 15 (StataCorp LLC, College Station, TX).

## RESULTS

Twenty-three women were screened for eligibility, and twelve were enrolled. Reasons for screen failure included current breastfeeding status, being more than 12 weeks past primary CIN2/3 treatment, among others. Participant baseline characteristics are summarized in Table 1. The mean age was 43.9 years (SD 4.4). Seven (58.3%) had a primary education or less as the highest level of education attained. Only 3 (25.0%) were formally employed, while the rest were farmers 4 (33.3%), self-employed 2 (16.7%), or unemployed 3 (25.0%). Five (41.7%) participants were married, and all 12 had a monthly income of less than KShs 25,000 (∼USD 120). All participants were on antiretroviral therapy with a median CD4 count of 781 cells/mm^3^ (IQR 418-890). Study uptake was 100.0% as every eligible participant offered enrollment agreed to enroll. All twelve participants started and completed all study visits. Nine (75.0%) of the participants elected to use the first dose of 5FU in the clinic under the observation of a study nurse. All participants used all eight 5FU doses per protocol. 5FU use was delayed for one week in one participant due to bothersome itching and burning sensation on urination associated with a Grade 1 AE.

**Table 1:** Characteristics of women with cervical intraepithelial neoplasia grade 2/3 (CIN2/3) who participated in th 5FU Phase 1 trial in Kisumu, Kenya (n=12)

| Characteristics | n (%) |
| --- | --- |
| **Age (years) Mean (SD)** | 43.9 (4.4) |
| **Highest education level attained** |  |
| Less than primary | 5 (41.7%) |
| Completed Primary | 2 (16.7%) |
| Completed Secondary | 4 (33.3%) |
| College or higher | 1 (8.3%) |
| **Occupation** |  |
| None | 3 (25.0%) |
| Salaried work | 3 (25.0%) |
| Business/Trader/Vendor | 2 (16.7%) |
| Farming | 4 (33.3%) |
| **Marital status** |  |
| Married/Living together | 5 (41.7%) |
| Divorced/Separated | 4 (33.3%) |
| Widowed | 3 (25.0%) |
| **Monthly income** |  |
| < Ksh 25,000 (\$200) | 12 (100.0%) |
| **Electricity available** |  |
| Yes | 5 (41.7%) |
| **Tap water available** |  |
| Yes | 5 (41.7%) |
| **Current or prior tobacco use** |  |
| No | 12 (100.0%) |
| **Parity Mean (SD)** | 3 (2) |
| **Age at first sexual intercourse (years) Mean (SD)** | 17.2 (2.1) |
| **No. of lifetime sexual partners Mean (SD)** | 4 (2) |
| **CD4 count** |  |
| Median (Q1, Q3) | 781.0 (418.0, 890.5) |
| **Currently contraception use** |  |
| Yes | 6 (50.0%) |
| No | 6 (50.0%) |
| **Time since HIV diagnosis (Years)** |  |
| 1-5 years | 2 (16.7%) |
| > 10 years | 10 (83.3%) |
| **Currently on ARVs** |  |
| Yes | 12 (100.0%) |
| **No. of years on ARVs** |  |
| 1 - 2 years | 1 (8.3%) |
| Greater than 2 years | 11 (91.7%) |
| **Lifetime cervical cancer screenings** |  |
| One | 7 (58.3%) |
| 2-3 | 3 (25.0%) |
| More than 3 | 2 (16.7%) |
| **Prior cervical precancer treatments** |  |
| One | 12 (100.0%) |
| **Precancer results prior to recent treatment** |  |
| CIN 2 | 2 (16.7%) |
| CIN 3 | 10 (83.3%) |
| **Precancer treatment received** |  |
| Thermal ablation | 5 (41.7%) |
| LEEP | 6 (50.0%) |
| Cryotherapy | 1 (8.3%) |

The type, frequency, and severity of participant-reported AEs are summarized in Table 2a. All 12 participants reported at least one grade I AE, 1 (8.3%) reported a grade 2 AE and no grade 3 or 4 AEs were reported. The most common AEs were grade 1 vaginal discharge in 9 (75.0%) and grade 1 pelvic pain in 7 (58.3%), with a mean duration of 3.2 and 2.1 days, respectively, all self-limited. One participant (8.3%) reported Grade 2 pelvic pain, which caused greater than minimal interference with daily activities and required non-narcotic medication. Of note, this participant reported pelvic pain at baseline (before 5FU initiation), which started after her excision procedure. Adverse events noted on pelvic exams during follow-up visits are listed in Table 2b. The most common was discharge in 8 (66.7%, 95% CI 34.9-90.0%) and cervical erythema 6 (50%, 95% CI 21.1-78.9%), all grade 1 (mild to moderate increase in discharge without pooling or erythema covering < 50% of the cervix). Four participants, 33.3% (95% CI 9.9-65.1%), had Grade 1 vaginal abrasions noted on exam, all superficial disruptions of the epithelium, measuring 0.5-1cm, with little to no mucosal involvement.

**Table 2a:** Type, frequency, and duration of participant-reported adverse events (AEs) among 5FU Phase I trial participants in Kisumu, Kenya (n=12)

| Reported symptoms | Grade 1<br>(n [%]) | Mean duration<br>days (SD) |
| --- | --- | --- |
| Abnormal vaginal discharge | 9 (75.0%) | 3.2 (1.7) |
| Pelvic pain | 8 (66.6%)<sup>1</sup> | 2.1 (1.3) |
| Vaginal irritation | 5 (41.6%) | 2.8 (1.2) |
| Headache | 4 (33.3%) | 2.3 (1.4) |
| Back pain | 3 (25.0%) | 2.8 (1.5) |
| Vaginal dryness | 2 (16.6%) | 2.3 (1.5) |
| Vaginal itch | 2 (16.6%) | 1.8 (0.96) |
| Fatigue | 2 (16.6%) | 3.5 (2.1) |
| Chills | 2 (16.6%) | 3.0 (2.8) |
| Painful urination | 1 (8.3%) | 1.3 (0.6) |
| Urinary frequency | 1 (8.3%) | 2 (0) |
| Fever | 1 (8.3%) | 5 (0) |
| Generalized itch | 1 (8.3%) | 2 (0) |
| Vulvar Irritation | 1 (8.3%) | 1 (0) |
| Myalgia | 1 (8.3%) | 1 (0) |

<sup>1</sup> 1 participant had a grade 2 pelvic pain, although this was present after her excision procedure and before starting 5FU use.. The pain caused greater than minimal interference with usual social and functional activities and treated with non-opioid analgesics.

**Table 2b:** Pelvic-exam identified adverse events (AEs) among 5FU Phase I trial participants in Kisumu, Kenya (n = 12)

| Observed symptoms | Grade | n (%) | 95% CI |
| --- | --- | --- | --- |
| Cervical erythema | 1 | 6 (50.0%) | (21.1% -78.9%) |
| Observed yellow discharge | 1 | 8 (66.7%) | (34.9% - 90.0%) |
| Vaginal abrasion | 1 | 4 (33.3%) | (9.9% - 65.1%) |
| Vulvar Erythema | 1 | 1 (8.3%) | (0.21% - 38.5%) |

All abrasions resolved with perineal care, which included the use of a peri bottle use during urination and the application of an occlusive moisturizer (Vaseline) to the affected area. One participant delayed the next dose of 5FU to ensure complete healing. No participant stopped 5FU use or did not complete the planned eight doses due to a lack of resolution of an abrasion. All participants tolerated all eight 5FU doses, and the majority of participants reported using tampons after each 5FU application. All 5 participants who had a male partner endorsed adherence to the recommendation of maintaining abstinence for two days after 5FU use.

On adherence assessment, evidence of 5FU use was supported by at least one of three methods for all 96 (100%) doses among the 12 participants. Four (4.2%) instances of 5FU use were not supported by one of three adherence assessment methods. In all four instances, the change in weight deviated slightly from what was expected, often a bit lower, and in two instances, the change in weight was higher than expected. In both cases, the participant endorsed spillage of cream when loading onto the applicator, and inspection of the returned applicator was consistent with streaking to the 2g mark, as expected. Hence, overuse was unlikely. Using all three methods to evaluate adherence, 96% of 5FU doses were used correctly. No participant was discontinued from the study because of non-compliance with the protocol.

## DISCUSSION

In this Phase I trial, adjuvant, self-administered, intravaginal 5%FU cream following primary CIN2/3 treatment among WLWH in Kenya was safe, tolerable, and associated with high adherence. All study participants tolerated all eight 5FU doses with no discontinuation due to primarily Grade 1AEs, and 96% of 5FU doses were used correctly based on three methods of adherence assessments. To our knowledge, this is the first study demonstrating these endpoints in a LMIC.

To achieve the WHO global cervical cancer elimination target of 90% of those with cervical precancer adequately treated by 2030, innovative, feasible, and scalable strategies are urgently needed in LMICs. Current precancer treatment gaps in LMICs include high precancer treatment failure rates among WLWH, as well as the inability of screen-positive women to access current provider-administered treatments. In Malawi, which has the world’s highest mortality from cervical cancer,^20^ between 2011 and 2015, only 43.3% and 31.8% of women with precancer who required cryotherapy or excision, respectively, received treatment.^21^ Similarly, in Kenya, a middle-income African country, linkage to treatment among those who screened position was below 40% between 2011-2020.^22^ Evidence of the efficacy of intravaginal 5FU as a primary cervical precancer treatment includes a randomized U.S trial where women with CIN2 had 93% regression rate at 6-months, compared to 56% randomized to observation (p<0.01). Such outcomes, if demonstrated in LMICs through repurposing 5FU as a self-administered primary precancer treatment, may be transformative in reaching millions of women unable to access provider-administered treatment and save millions of lives.

Self-administered intravaginal 5FU as adjuvant therapy following primary treatment could also improve precancer treatment outcomes in WLWH in LMICs who face 25-30% CIN2/3 recurrence after treatment.^4,7,23^ A U.S. randomized trial of self-administered 5FU as adjuvant therapy after CIN2/3 treatment among WLWH using the same 5FU protocol as our study demonstrated 8% recurrence at 18 months, compared to 31% in the observational arm.^12^ Currently, there is a randomized trial on the feasibility of self-administered intravaginal 5FU after LEEP among South African WLWH (Clinicaltrials.gov NCT05413811).

Early 5FU treatment regimens were associated with severe side effects, including chronic vaginal ulcerations, often associated with daily or twice daily application for multiple days at a time,^24^ which deterred its use. However, the recent studies described above limiting 5FU use to less frequent applications demonstrate an acceptable side effect profile, consistent with our findings. In our study, the most notable AE was minor vaginal abrasions noted on the exam on 4 (33.3%, 95% CI 9.9-65.1%) participants. These were all grade 1 superficial epithelial disruptions with little to no mucosal involvement, which were self-limited and managed conservatively with a peri bottle to reduce discomfort with urination and a topical occlusive barrier (Vaseline) for comfort. In one participant, the next 5FU dose was delayed by one week to ensure healing. In a recent U.S feasibility trial of alternating 5FU and imiquimod for primary CIN2/3 treatment, 2 (15.4%) had Grade 1 and 2 (15.4%) Grade II vaginal abrasions, defined as size <1cm, all of which resolved spontaneously.^25^

Our findings of a slightly higher rate of vaginal abrasions may be related to our study population being older and perimenopausal (mean age of 43.9 years (SD 4.4)), compared to a mean age of 27 years in the U.S. study. It is reassuring that all abrasions in our study were grade 1, self-limited, and did not interfere with daily activities. Similar to a recent study of intravaginal 5FU for vaginal precancer,^11^ we recommended the use of an occlusive emollient cream to the external genitalia before and for 2-3 days after 5FU use for barrier protection. Larger studies of self-administered 5FU in LMICs can investigate the frequency of this AE, as well as its impact, if any, on HIV genital shedding among users. Future studies can investigate whether a lower dose (e.g., 1g instead of 2g) or fewer applications may reduce abrasions, particularly in menopausal women.

This study’s strengths are its use of a standardized scale to evaluate safety, the frequency of study visits (every two weeks), and no loss-to-follow-up, limiting ascertainment bias. Additionally, adherence was evaluated using three different methods, increasing validity. Limitations include a small sample size and being a single-site study, which may reduce generalizability. Future larger from multiple sites can address these limitations. Future studies can evaluate the acceptability of these therapies among study participants, their partners, and healthcare stakeholders.

In summary, this study demonstrates that self-administered intravaginal 5FU, as adjuvant therapy among African women with CIN2/3 following primary treatment, was safe, well tolerated, and associated with very high adherence rates. Over half of the participants in our study had a primary education or less, and the majority had no electricity or tap water in their homes. Our finding of high safety and adherence in this population suggests that topical therapies like 5FU can be safely used in rural African settings, where access to provider-administered precancer therapy is limited. Randomized efficacy trials are needed to investigate whether adjuvant 5FU can improve precancer treatment outcomes among WLWH in LMICs or be used as a primary therapy to address current gaps. An effective self-administered topical therapy for cervical precancer in LMICs may be transformative in increasing treatment access and save millions of lives.

## AUTHOR CONTRIBUTIONS

CM: conceptualization, data analysis and interpretation, drafting, final approval, JO: conceptualization, data interpretation, final approval, CO: study implementation, data collection, final approval, AO: study implementation, data collection, final approval, MR: study implementation, data collection and analysis, final approval, GR: study implementation, data collection, final approval, EA: conceptualization, data interpretation, final approval, JT: conceptualization, data interpretation, critical revisions, final approval, LR: conceptualization, data interpretation, critical revisions.

## FUNDING

This research was supported by the Women’s Reproductive Health Research Career Development Grants (K12 HD10308) and the Victoria’s Secret Global Fund for Women’s Cancers Career Development Award, in Partnership with Pelotonia and the American Association of Cancer Research (AACR).

## CONFLICT OF INTEREST

The authors declare no conflict of interest.

## DATA AVAILABILITY STATEMENT

Anonymized research data are available upon reasonable request.

---

**Supplemental Image 1:**
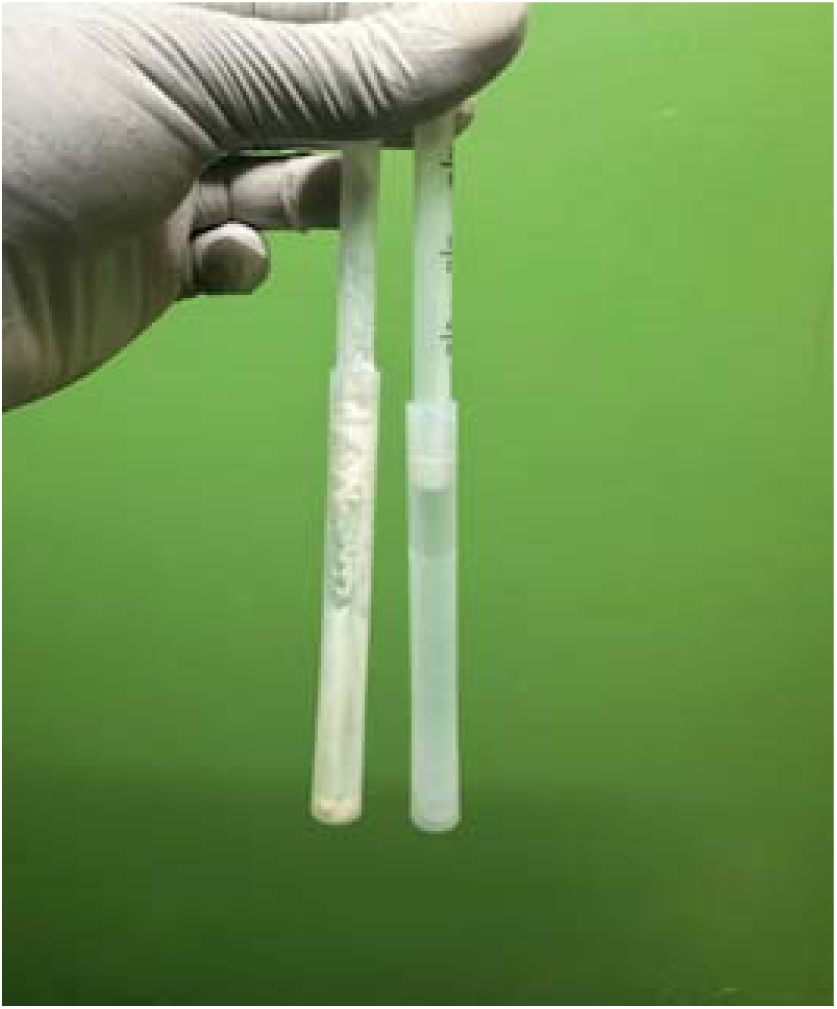
Example image of inspection of a returned vaginal applicator for evidence of exposure to the study drug as part of adherence assessment Used applicator on the left compared to unused applicator on the right

## Notes

### Competing Interest Statement

The authors have declared no competing interest.

### Clinical Trial

NCT05362955

### Funding Statement

This research was supported by the Womens Reproductive Health Research Career Development Grants (K12 HD10308) and the Victorias Secret Global Fund for Womens Cancers Career Development Award, in Partnership with Pelotonia and the American Association of Cancer Research (AACR).

### Author Declarations

The study was approved by the institutional review boards at the University of North Carolina at Chapel Hill (21-3265) and the Kenya Medical Research Institute (KEMRI/SERU/CMR/4555). All participants provided informed consent.

